# Acoustic Epidemiology of Pulmonary Tuberculosis (TB) & Covid19 leveraging AI/ML

**DOI:** 10.1101/2022.02.05.22269707

**Authors:** Rahul Pathri, Shekhar Jha, Samarth Tandon, Suryakanth GangaShetty

## Abstract

Involuntary cough is a prominent symptom for many a Lung Ailments ranging from Infectious to non-Infectious diseases. Early research around human cough established that the spectral signatures do not vary between Involuntary and Voluntary coughs. The study aimed at evaluating voluntary human cough sounds recorded under a stringent clinical protocol. India’s ambitious goal to eliminate and eradicate TB by 2025 shall be facilitated by Machine Learning tools that address subjectivity in that the healthcare worker can now take the solution as a screening modality to the last mile as a part of outreach programs without having to rely on infrastructure & connectivity. In this paper we present the findings of Clinical Trials for Pulmonary TB registered at CTRI/2019/02/017672 conducted independently and included Covid19 during the pandemic as a part of Bi-Directional screening modality. The reference standards used were CBNAAT (Cartridge based nucleic acid amplification test) & CXR (Chest X-Ray) for TB while for Covid19; RT-PCR was used as the reference standard. As a non-invasive and contactless screening modality, a sophisticated third party Microphone Array was used to record the cough under a stringent infection control protocol. Sensitivity achieved across the sites for TB ranged between 80% - 83% and Specificity was to the tune of 92% while using CBNAAT as a reference standard. CXR when used as a reference standard for TB achieved a sensitivity and specificity of 59% and 60% respectively. Covid19 achieved a sensitivity & specificity of 92% and 96% while using RT-PCR as the reference standard. The study was primarily focused on the Frequency domain that paved way for feature extraction and explainable Machine Learning Models operating upon lossless WAV files hypothesizing acoustic theory and demographic inputs. The solution titled “TimBre” can now be added to the healthcare workers arsenal in situations where a RT-PCR or CXR is not available and seamlessly conduct bi-directional screening with a single recording of cough and also offer insights into Non-Communicable diseases as a part of differential diagnosis.

## 1) Introduction

One of the famous quotes by Albert Einstein around music was – “It occurred to me by intuition, and music was the driving force behind that intuition. My discovery was the result of that musical perception” - (0).

Human Lungs infected with TB or Covid19/Pneumonia introduces a certain Lung parenchymal changes that occasionally overlaps or mimics other Lung conditions. In case of TB, these are Cavities at different stages & in case of Pneumonia, alveoli getting filled with puss and other liquids forming liquefaction followed by air/fluid filled cavities(1). These changes may be hypothesized with an Acoustic Guitar and other accessories that are discussed in this paper. Pulmonary Tuberculosis share of the global TB burden is to the tune of 70% with remaining cases being extra pulmonary & the numbers are increasing due to the obscuring of TB care as a result of the Pandemic caused by Covid19. In this paper we present findings of a non-invasive, point of care & real time results driven solution that addresses screening of both TB and Covid19 affecting the Lungs as a potential to be a bidirectional screening modality.

## 2) Materials and Methods

Informed consent was obtained via a traditional method to obtain signatures on paper available in English, Kannada and Bengali languages for Site2 that was conducted at NH, Bangalore while subsequent sites leveraged the digital screen on the TimBre app that is available as a private APK as of today. The app is not available on Google Playstore given the fact that without a Microphone Array, the results could get compromised and cause unnecessary anxiety among users. Across the sites a very stringent hygiene protocol was mandated wherein the healthcare worker was equipped with n-95 masks, gloves while the subject coughing had to wear a surgical mask provided by the PI. The 3^rd^ party microphone array filters were replaced after each cough and incinerated. The Microphone Array used was zoom H1 and H1n models and a representative image with the filter and is depicted in the following image:

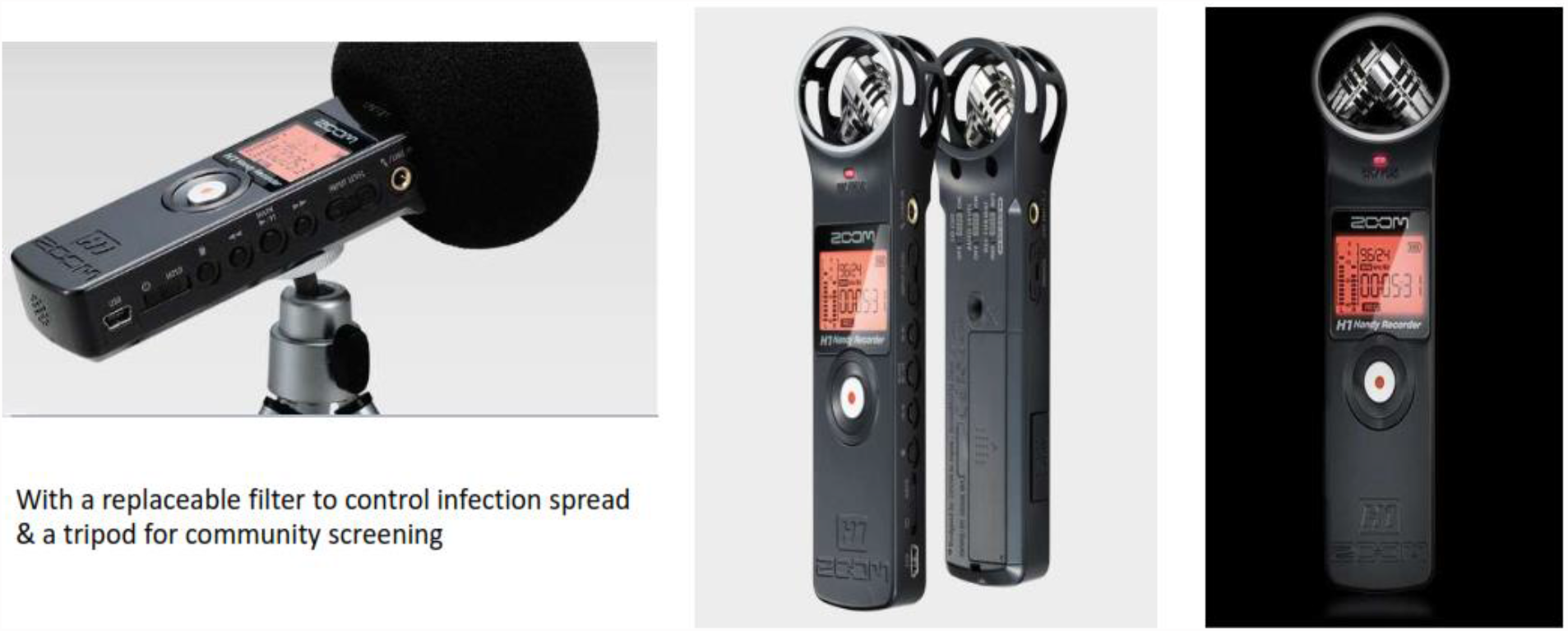

The array was connected to the Mobile phone USB port via an OTG cable & subjects were advised to take a deep breath before coughing onto the array with a filter for approximately 10 seconds while the distance between the mouth and the device was approximately at an arm’s length divided by 2. The deep breath ensured no superficial coughs were recorded. The TimBre app would also obtain demographic information pertaining to height and weight that would get recorded by a status meter and a digital weighing scale respectively for subsequent arrival of the BMI given the fact that known TB cases in the Machine Learning Model had a BMI lower that 18 for most part unless they were recovering. To maintain consistency with the Machine Learning Model data, clinical trial sites recorded cough sounds at 44.1 kHz for both TB and Covid19. The feature extraction resulted in data for both Left and Right channels facilitated by the XY design of the microphone array. While the array had built in ability to eliminate undesirable noise via a low cut filter & other settings, we also applied background noise reduction techniques that would eliminate any conversations between the healthcare worker and the subject. The TimBre app would facilitate data transfer of the WAV file and demographics to the cloud (Azure, IBM, GCP, and AWS) for further processing that included the following order:

1. Feature Extraction: Extracting features from both the channels for various frequency bands ranging from 0-200 Hz till 5000 Hz with an interval of 500 Hz for each band starting from 500Hz. The features extracted for both the XY channels were – Sum, Standard Deviation, Variance, Coefficient of Variance, Top10 Amplitude, Spectral Centroid, Spectral Flatness, Spectral Skewness, Kurtosis, MFCC and Energy
2. Compute the BMI and append height, weight & BMI to the feature extracted spectral record
3. Score against a RUS (random under sampling) Boosted Model given imbalanced class sets for TB and Covid19. Most importantly the advantages of Ensemble Models as seen from the higher AUC (Area under curve) & Sensitivity/Specificity depicted by the by the ROC curve (receiver operating characteristic)
4. Shared the results with an SLA of 48 hours on a secured portal that would also generate a unique patient id
5. Results were classified as 0-Negative, 1-Positive and 2-Rejected due to lack of fidelity in the sound file

Spectrograms were not considered due to the fact that the MFCC (mel frequency cepestral coefficients) components were already included as a part of feature extraction & the sheer volume of data required for Deep Learning models was not available with an added risk of misrepresentation of data when augmented using tools & techniques such as VTLP – vocal tract length perturbation or MATLAB augmentation techniques. Most importantly, lack of explainability & interpretability with deep learning models

Starting from 2018 several pilot studies were conducted at schools, PLHIV, colleges, factories, courts, home for the aged and these were primarily for data harvesting for AI/ML models. The below table depicts the actual pre-clinical & clinical trial related studies:

**Table 1:**
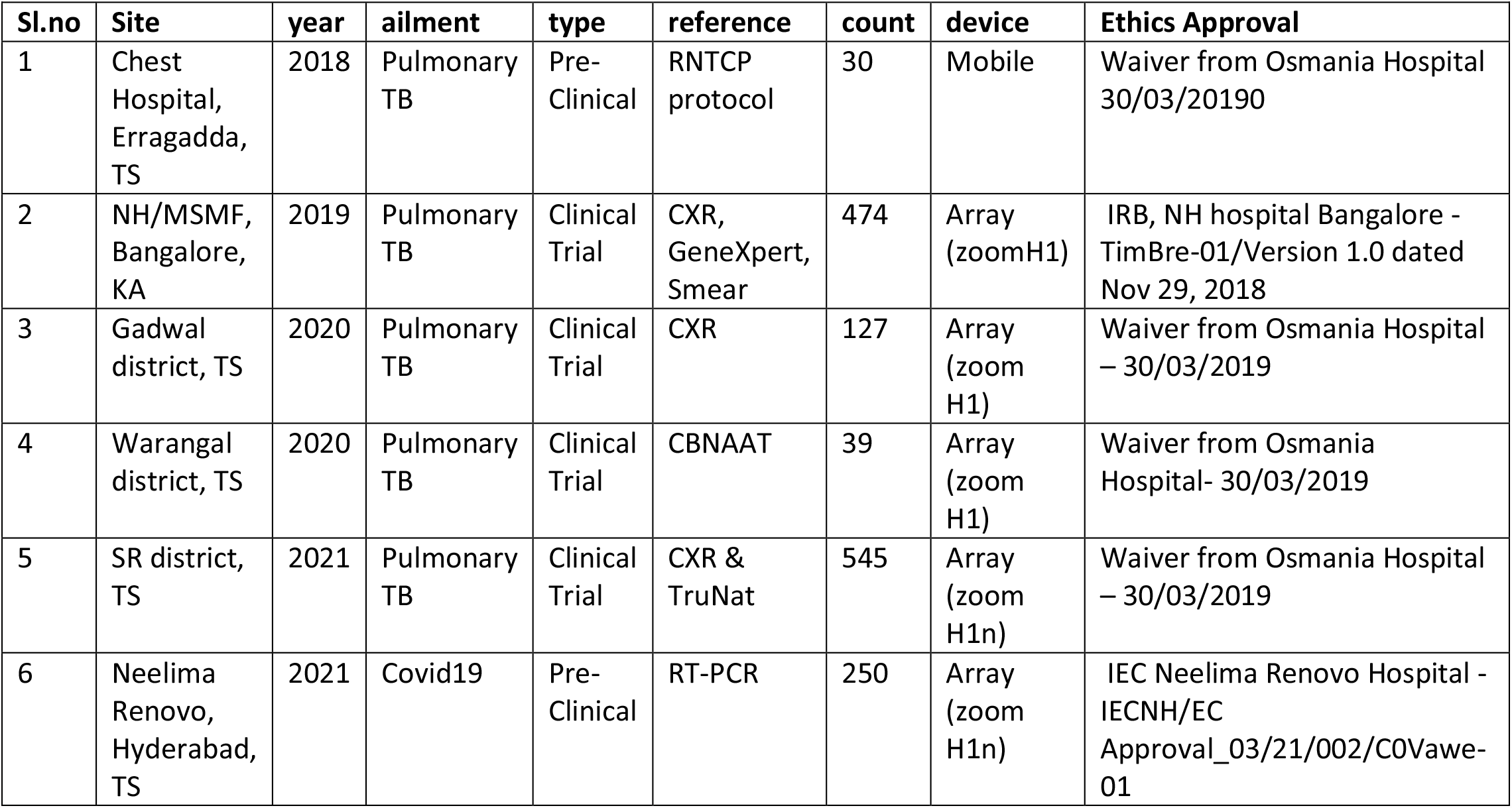
Clinical Trials and Pre-Clinical Validation.

| Sl.no | Site | year | ailment | type | reference | count | device | Ethics Approval |
| --- | --- | --- | --- | --- | --- | --- | --- | --- |
| 1 | Chest Hospital, Erragadda, TS | 2018 | Pulmonary TB | Pre-Clinical | RNTCP protocol | 30 | Mobile | Waiver from Osmania Hospital 30/03/20190 |
| 2 | NH/MSMF, Bangalore, KA | 2019 | Pulmonary TB | Clinical Trial | CXR, GeneXpert, Smear | 474 | Array (zoomH1) | IRB, NH hospital Bangalore - TimBre-01/Version 1.0 dated Nov 29, 2018 |
| 3 | Gadwal district, TS | 2020 | Pulmonary TB | Clinical Trial | CXR | 127 | Array (zoom H1) | Waiver from Osmania Hospital – 30/03/2019 |
| 4 | Warangal district, TS | 2020 | Pulmonary TB | Clinical Trial | CBNAAT | 39 | Array (zoom H1) | Waiver from Osmania Hospital- 30/03/2019 |
| 5 | SR district, TS | 2021 | Pulmonary TB | Clinical Trial | CXR & TruNat | 545 | Array (zoom H1n) | Waiver from Osmania Hospital – 30/03/2019 |
| 6 | Neelima Renovo, Hyderabad, TS | 2021 | Covid19 | Pre-Clinical | RT-PCR | 250 | Array (zoom H1n) | IEC Neelima Renovo Hospital - IECNH/EC Approval_03/21/002/C0Vawe-01 |

Zoom H1n has certain built-in features to source power from the Mobile phone, auto level, limiter that were leveraged. The streaming feature introduced in the latest model however gave inferior results & hence we continued to use WAV files recorded and stored on the SDCard as opposed to direct streaming of WAV files onto the cloud. Since the existing Machine Learning models harvested WAV files at a 44.1kHz / 16 bit, we continued with the SDCard approach which is considered as a gold standard.

## 3) Clinical Trial Results & Statistics

### Sl.no 1 Table1

Preclinical validation was conducted on known data sets & golden truth was obtained from the case sheets of the Government Chest Hospital. At the time of running the Algorithms for prediction, the results were blinded. Study was broken into 5 cohorts & the results of SCORED data are below wherein the numerator represents the results of the Algorithm and the denominator represents the cohort count.

1– Healthy Patients (10/10)

2- MDR-TB (4/5)

3- Old PTB (3/5)

4- EPTB (Pleural Effusion) (2/3)

5- PTB (5/5)

One MDR (multidrug drug resistant) was misdiagnosed and one Pleural Effusion was diagnosed as false positive (needs clinical evaluation) since the patient may still be sputum positive. All healthy cases are diagnosed as negative & all known PTB are diagnosed as positive. The old TB patient’s diagnosis was unavailable and hence we labelled positive for anyone with a BMI lower than 17.5.

### Data Collection & Machine Learning

#### Screenshot -1

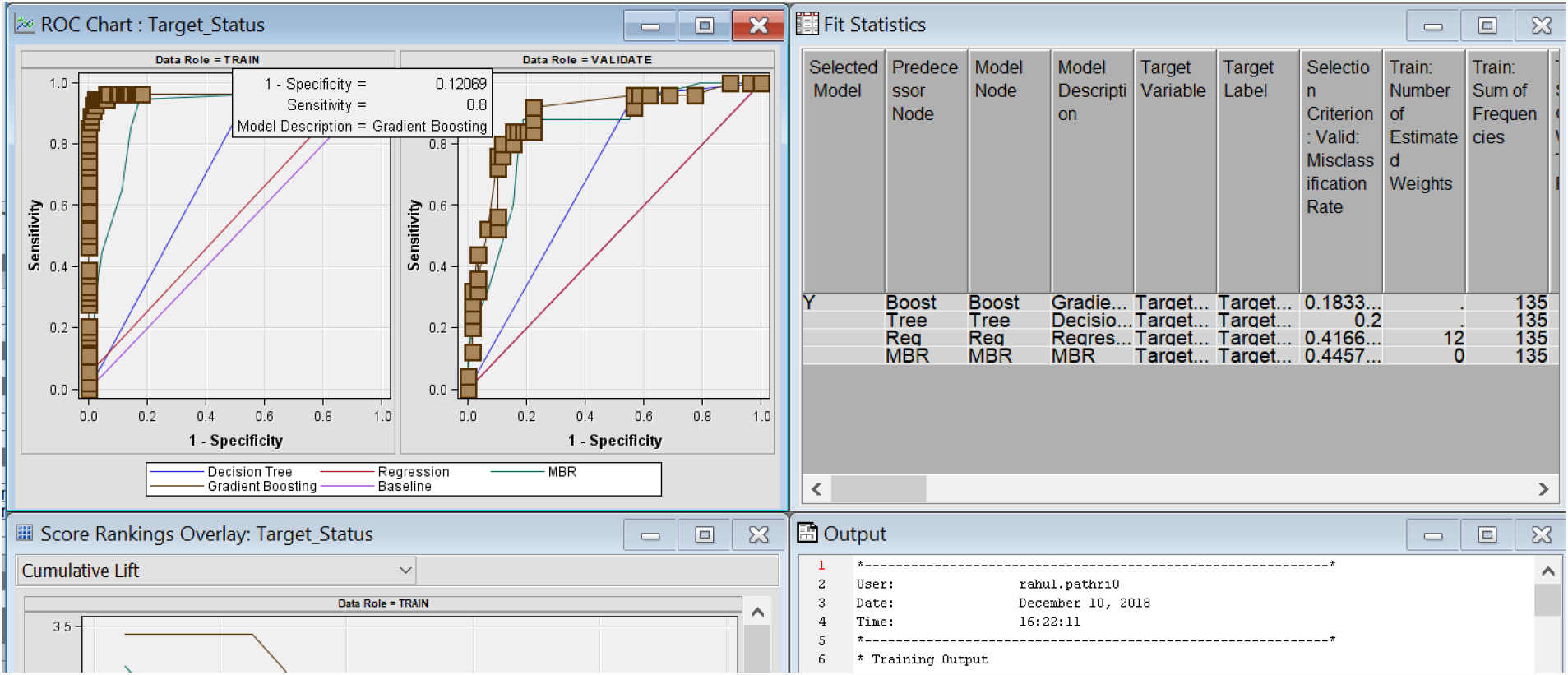

➢ Data was collected using a Xaomi Redmi 6a smart phone for all the patients across three different devices by our field staff after obtaining patient consent
➢ Instrument was cleaned after each cough and made the patient wear a surgical mask while coughing into the phone
➢ The device was equipped with a single channel built-in microphone
➢ Data was partitioned into 70% Training and 30% Validation for a record set of 250 Patient
➢ Sensitivity was reported as 80% and specificity as 88% as seen from the ROC curve – screenshot-1
➢ Gradient Boosting was the Algorithm chosen with the lowest misclassification rate – Screenshot-1

### Sl.no 2 Table1

The study was conducted at NH/MSMF hospital, Bangalore and the cohort included non-suspicious cases that were asymptomatic. Data was collected by the hospital staff and all of the activity was double blinded. Results were shared with the PI within 48 hours

#### Clinical Trial Statistics

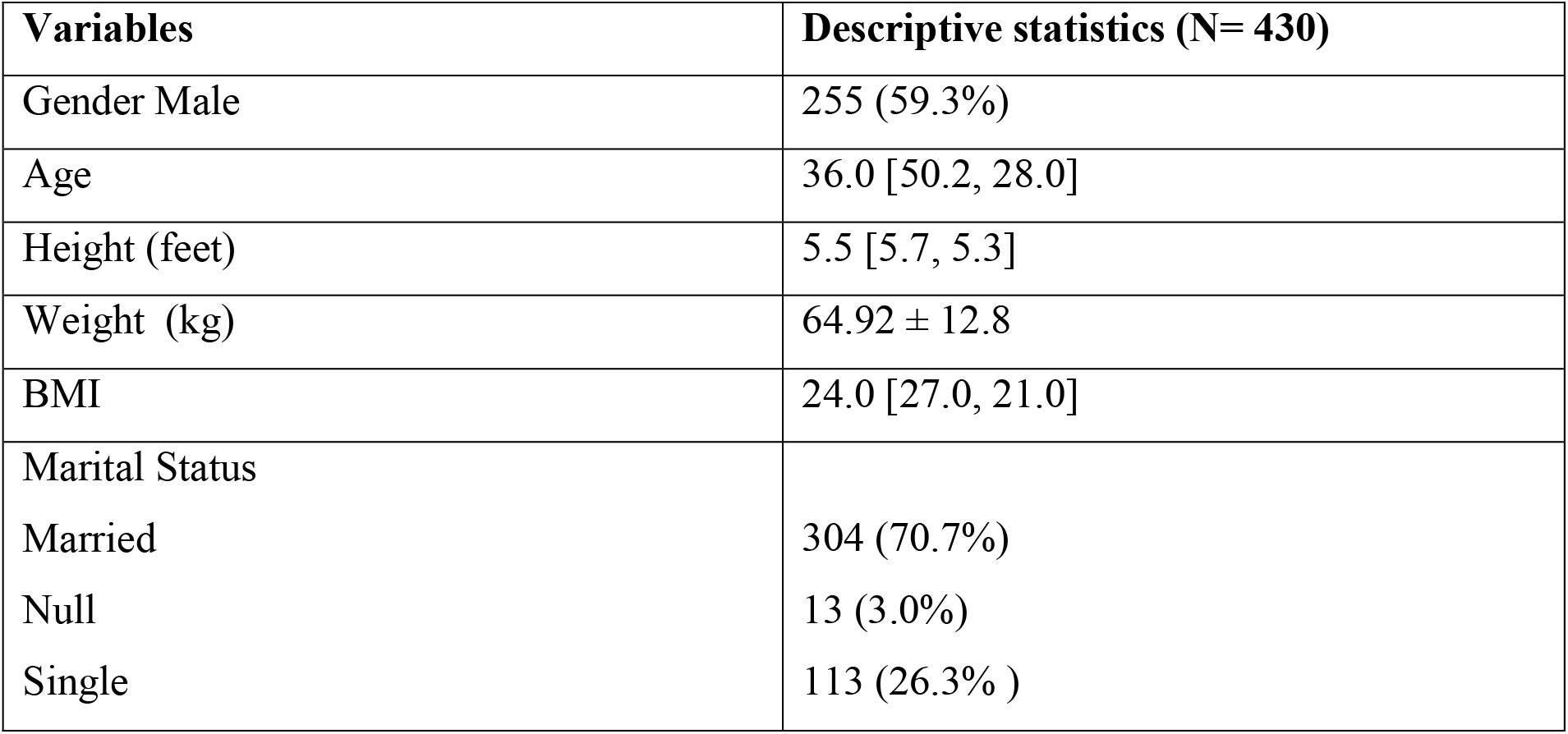
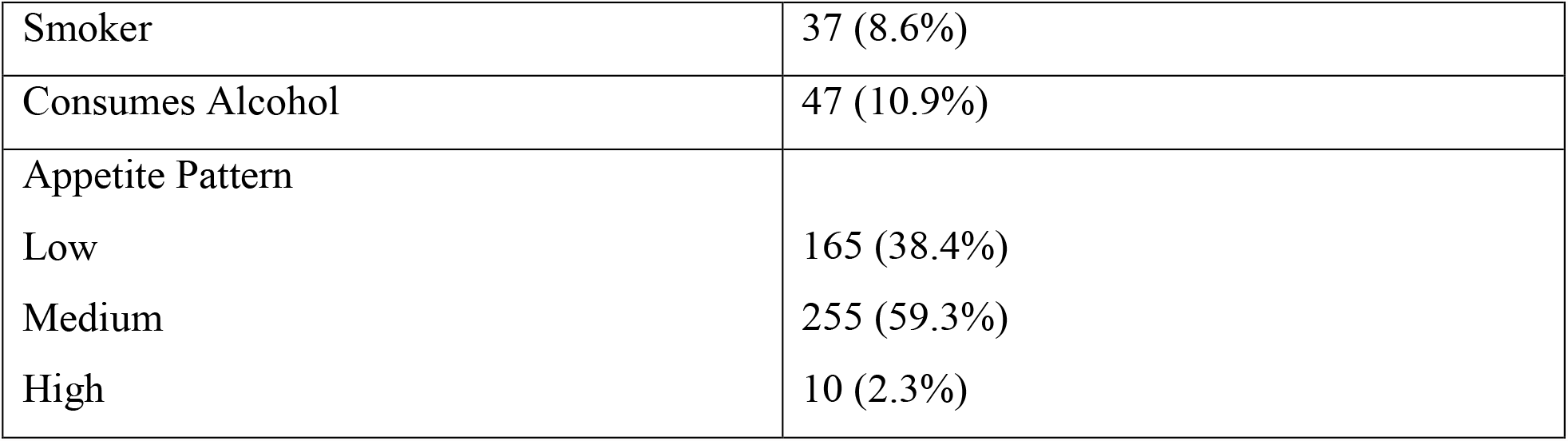
**Results: Group 2 Table-1 Baseline characteristics**

A total of 474 patients were recruited as of Feb 8, 2020. Among them 39 patients were diagnosed as positive by TimBre software. The details of their follow up are mentioned as follows:

**Table 2.**
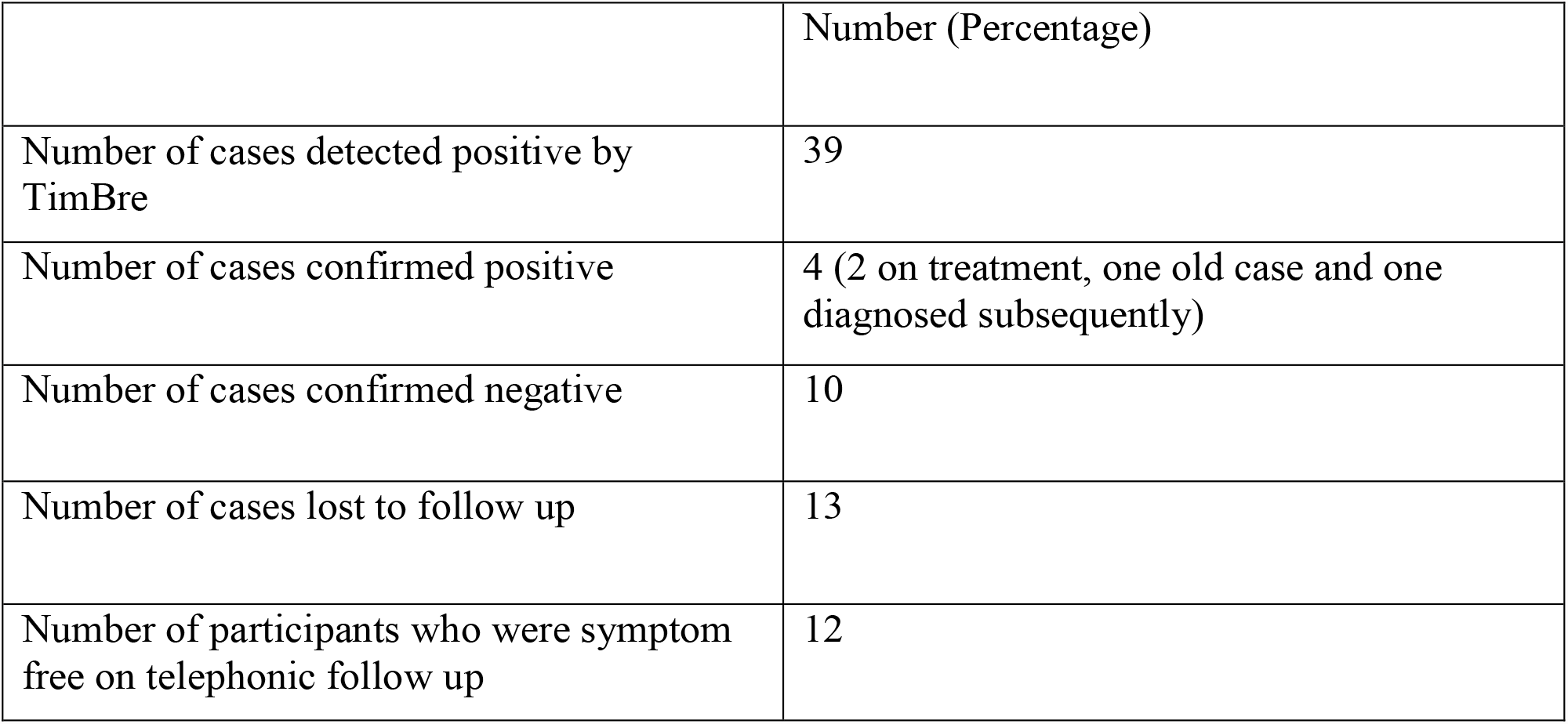
TimBre results for 474 patients.

#### Number of cases missed diagnosis of TB by TimBre 1

For a total of 474 participants screened 39 were detected as positive by TimBre software. Among the 39 participants 4 were positive. Two were on ATT at the time of testing for TimBre software. One was an old case with no active bacilli in sputum. One patient was later found to be positive on further evaluation.

Ten participants had clinical evaluation and were confirmed as negative for TB by either clinical examination or sputum testing. Thirteen patients could not be reached further for telephonic follow up or hospital visit. Remaining 12 patients were followed telephonically for symptoms suggestive of TB like chronic cough, fever and weight loss. None of these 12 patients were symptomatically positive for tuberculosis.

One patient who was positive by GenXpert was missed by the software.

**Table 3.**
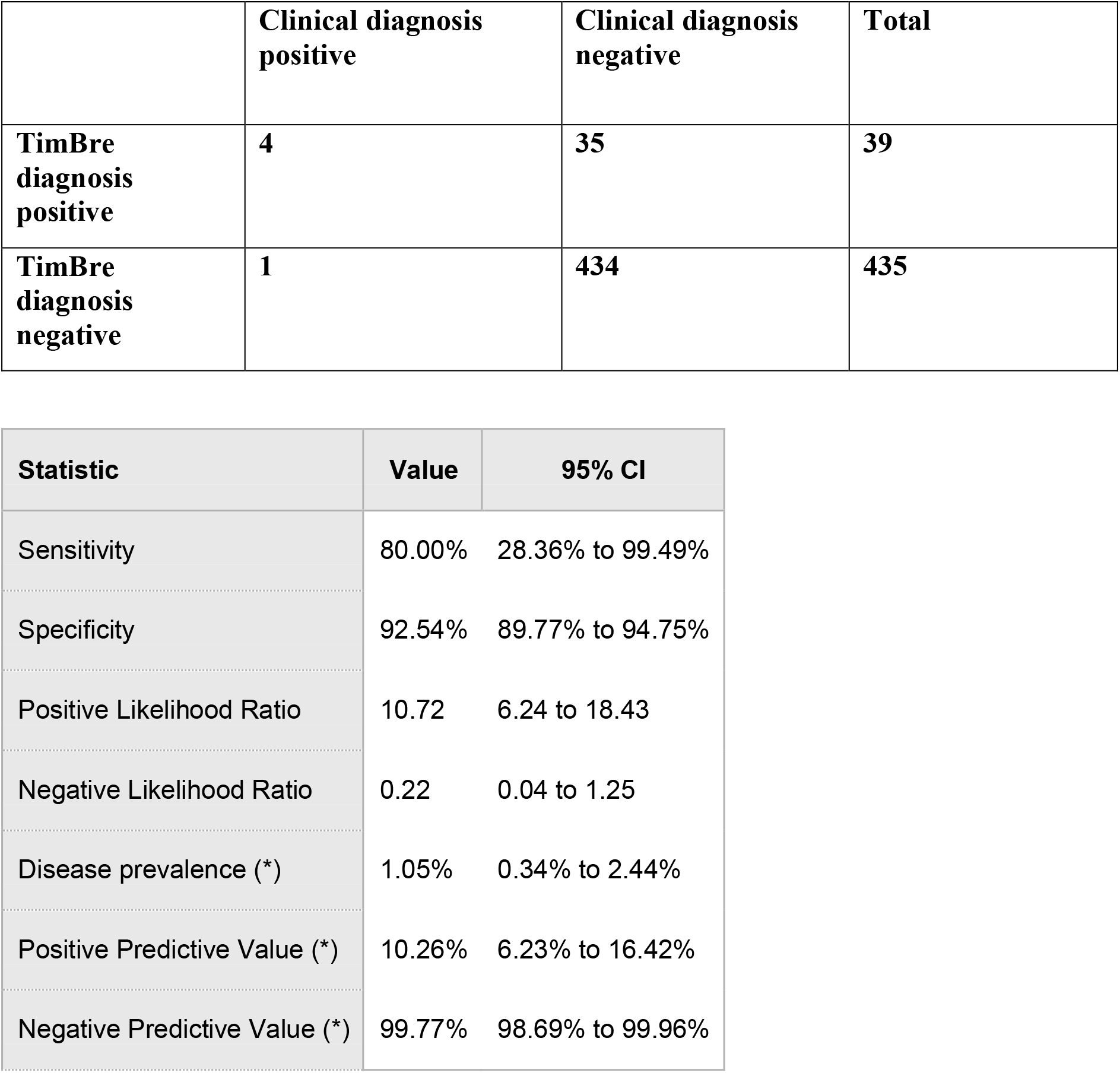
Sensitivity, specificity of TimBre.

### Sl.no 3 & 4 Table1

#### Clinical Trial Statistics

##### Site-1 Warangal district

**Table 1.**
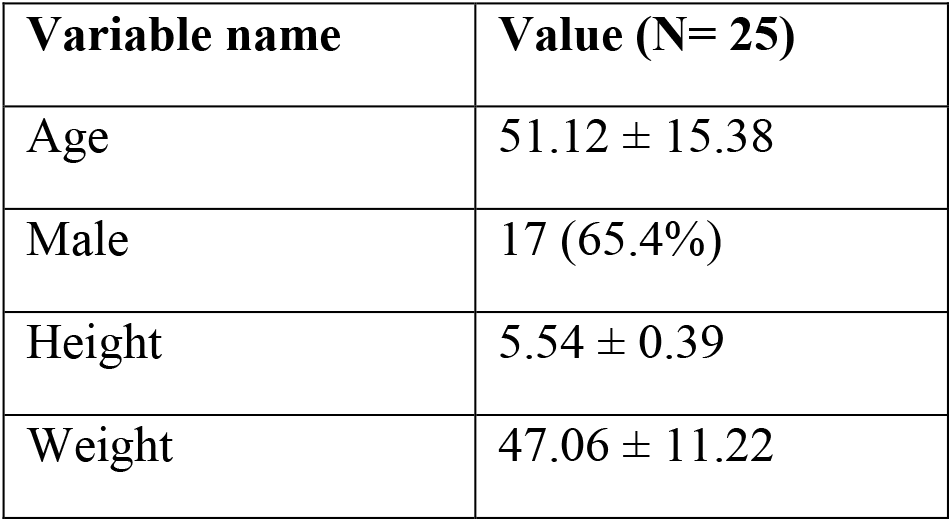
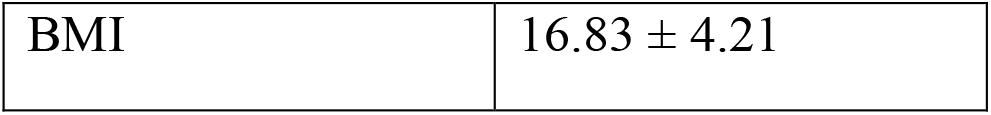
Baseline demographic characteristics.

**Table 2.**
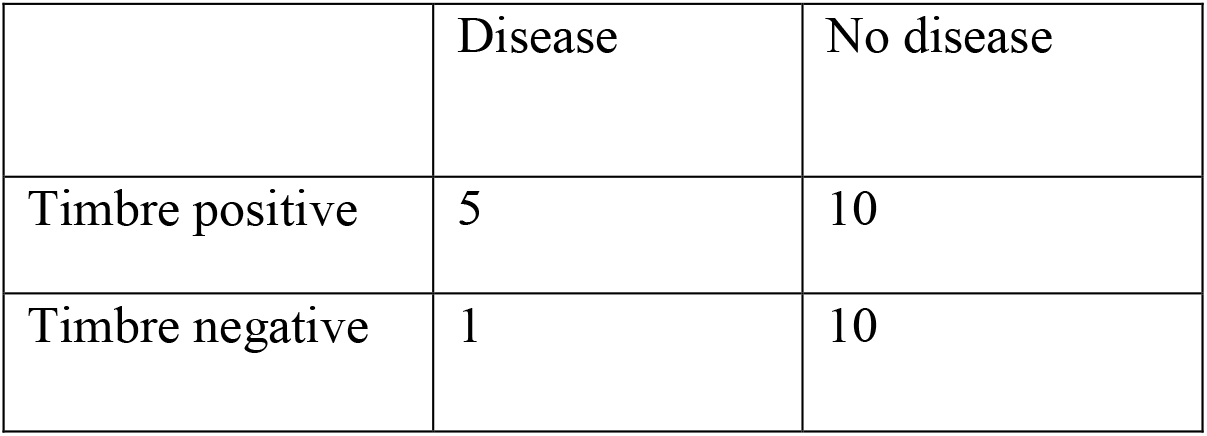
Comparison of TimBre results with chest X-ray.

**Table 3.**
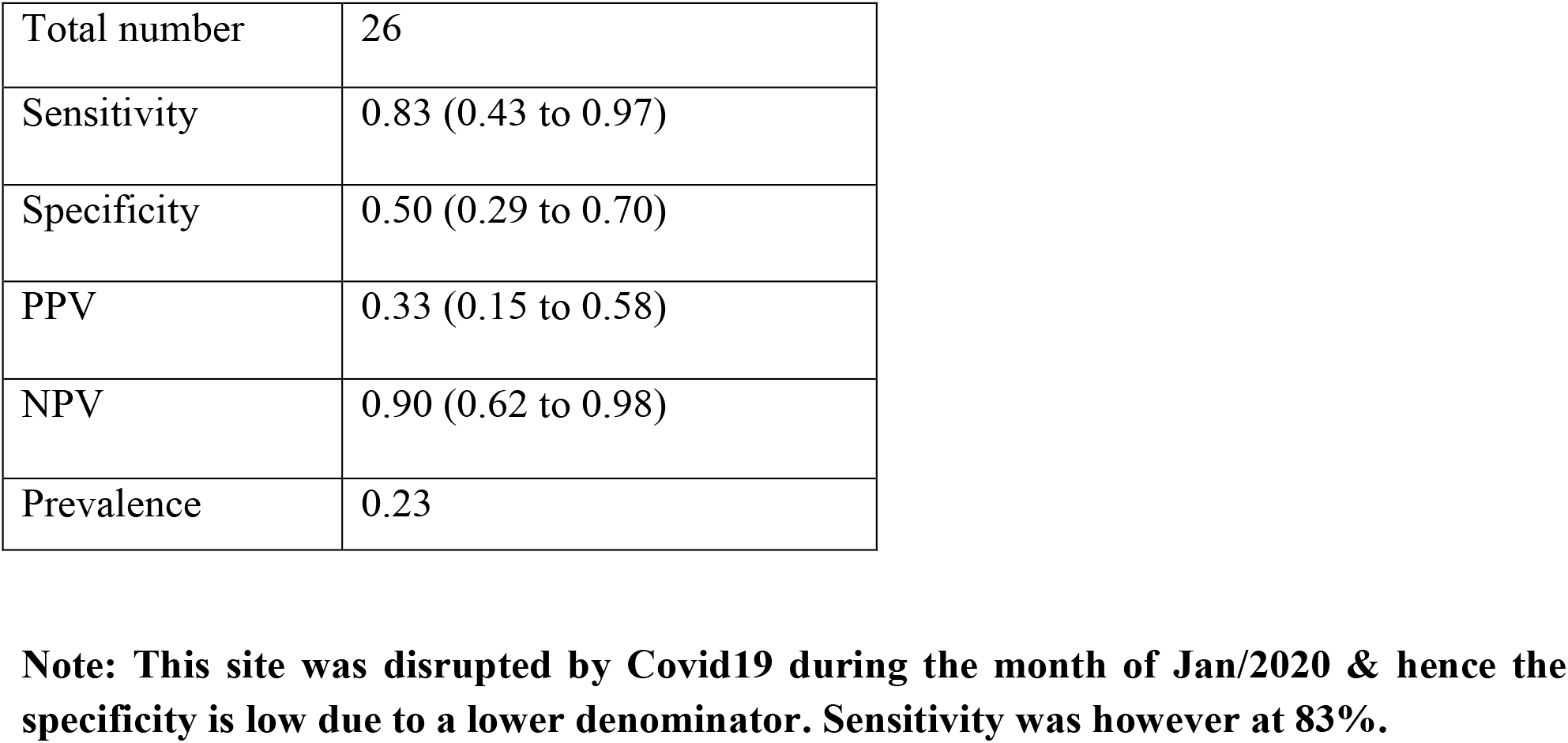
Diagnostic accuracy of TimBre.

|  |  |
| --- | --- |
| Total number | 26 |
| Sensitivity | 0.83 (0.43 to 0.97) |
| Specificity | 0.50 (0.29 to 0.70) |
| PPV | 0.33 (0.15 to 0.58) |
| NPV | 0.90 (0.62 to 0.98) |
| Prevalence | 0.23 |

**Note: This site was disrupted by Covid19 during the month of Jan/2020 & hence the specificity is low due to a lower denominator. Sensitivity was however at 83%.**

##### Site-2 Gadwal district

**Table 4.** Baseline demographic characteristics.

**Site-2 Gadwal district** **Table-4 Baseline demographic characteristics**
| Variable name | Value (N= 125) |
| --- | --- |
| Age | 51.41 ± 15.64 |
| Male | 83 (66.93%) |
| Height | 5.20 ± 0.34 |
| Weight | 45.78 ± 10.81 |
| BMI | 18.70 ± 4.15 |
| Smoking | 21 (16.93%) |
| Appetite<br>Low<br>Medium | 89 (71.77%)<br>35 (28.22%) |
| Alcohol | 23 (18.54%) |

**Table 5.**
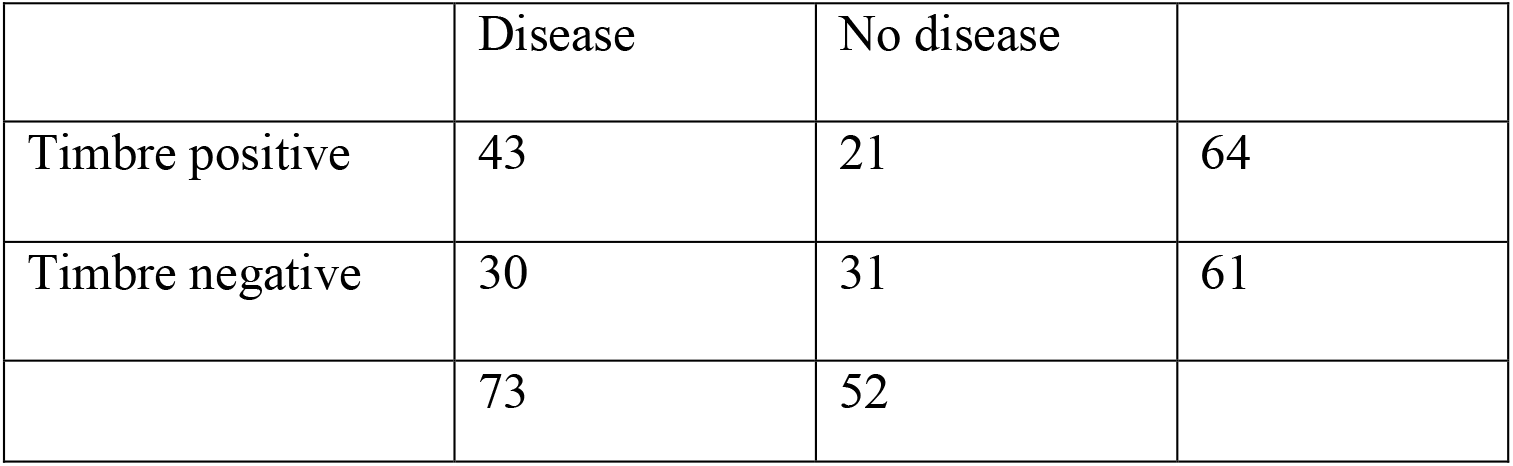
Comparison of TimBre results with chest X-ray.

|  | Disease | No disease |  |
| --- | --- | --- | --- |
| Timbre positive | 43 | 21 | 64 |
| Timbre negative | 30 | 31 | 61 |
|  | 73 | 52 |  |

**Table 6.**
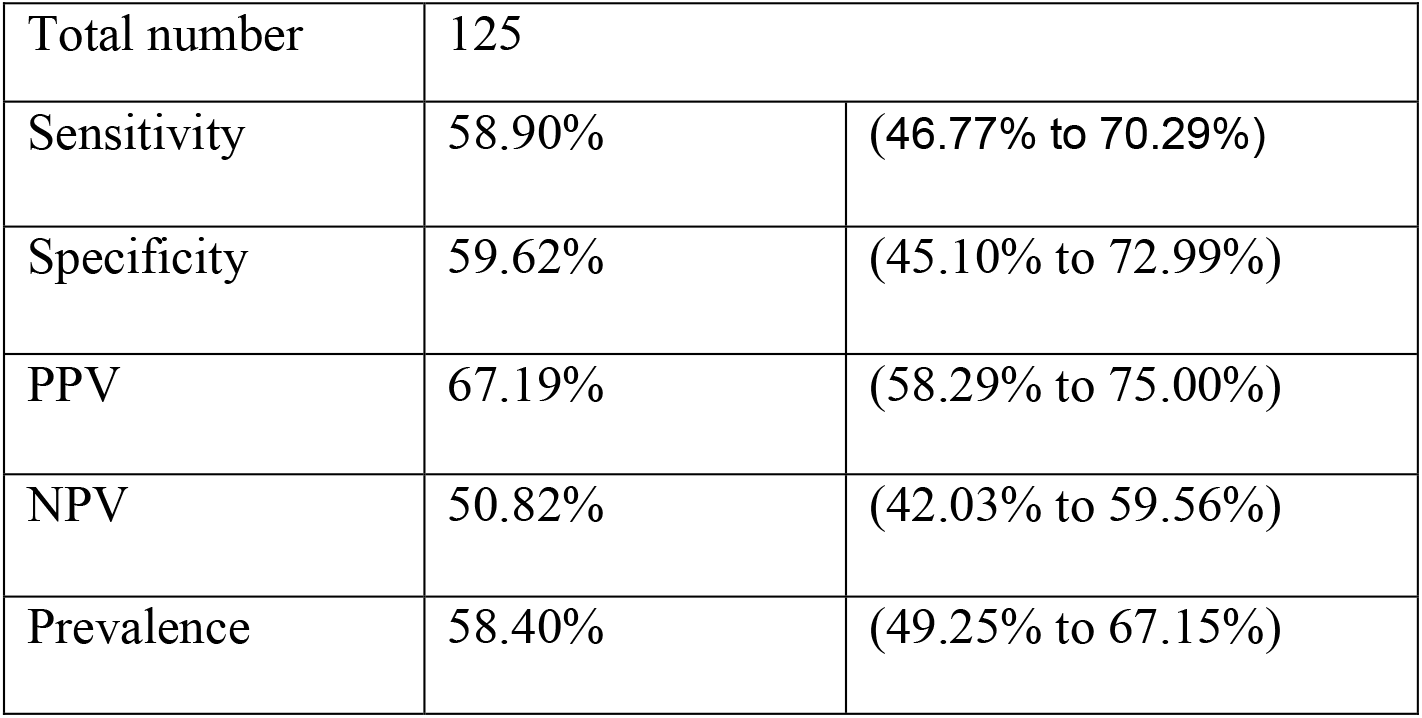
Diagnostic accuracy of TimBre.

### Sl.no 5 Table1

This site was the first site to adopt bidirectional screening & used the reference standard CXR and TruNat. Only 77 out of 545 results were available and remaining were lost to follow up. The concordance with CXR was found to be 100% and 25% with TruNat. A glitch found on the field during the trial was related to streaming of the WAV files and also using C-Type USB convertor connecting the array to latest Oppo phone models cascaded the glitch & delayed reporting. A total of 57 presumptive positive Covid19 were reported that were not subjected to the reference standard RT-PCR confirmatory test due to constraints caused by the Pandemic and also focus being on the Active Case Finding (ACF) for TB cases. The Machine Learning models AUC was at 92% with a sensitivity and specificity of 81% and 92% respectively for the RUS Boosted Models built separately for both TB and Covid19. We leveraged existing Pneumonia data for Covid19 models. Across the sites (Sl.No-2,3,4,5 & 6 of Table-1), a 10-Fold cross validation was used to avoid over-fitting that implied the number of partitions facilitating each record to have an equal chance of being both in the Training and Validation sets as a part of the Ensemble Model

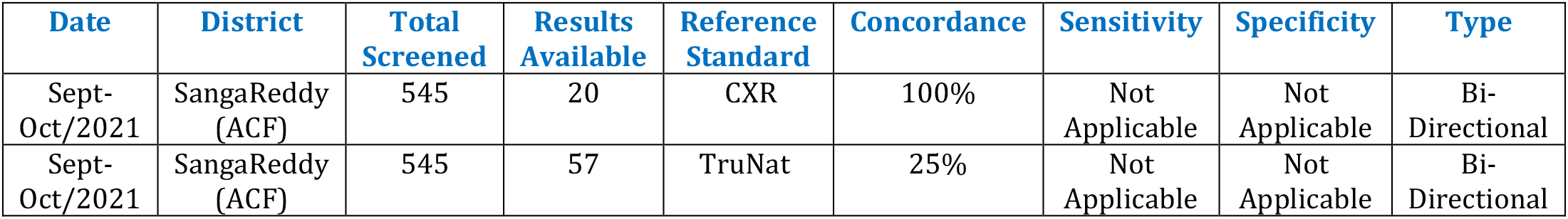

| Date | District | Total Screened | Results Available | Reference Standard | Concordance | Sensitivity | Specificity | Type |
| --- | --- | --- | --- | --- | --- | --- | --- | --- |
| Sept-Oct/2021 | SangaReddy (ACF) | 545 | 20 | CXR | 100% | Not Applicable | Not Applicable | Bi-Directional |
| Sept-Oct/2021 | SangaReddy (ACF) | 545 | 57 | TruNat | 25% | Not Applicable | Not Applicable | Bi-Directional |

### Sl.no 6 Table1

The Covid19 preclinical validation used RT-PCR as a reference standard and the golden truth was known during data harvesting and the results were blinded to the Algorithm during prediction and achieved at sensitivity & specificity of 92% and 96% respectively for a total screened patient count of 250 that included both Mobile Screening using Nokia 2.4 and the Microphone Array. The PPV and NPV were at 97% and 88% respectively

## Discussion & Conclusion

### Screenshot-3

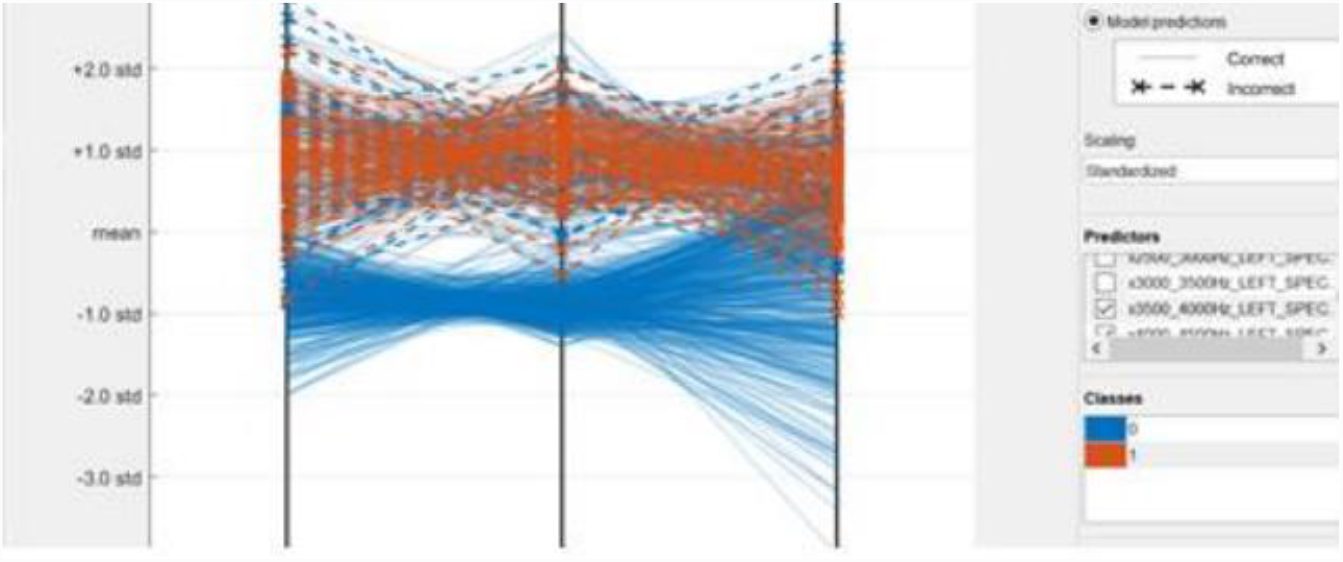

**Table-3.**
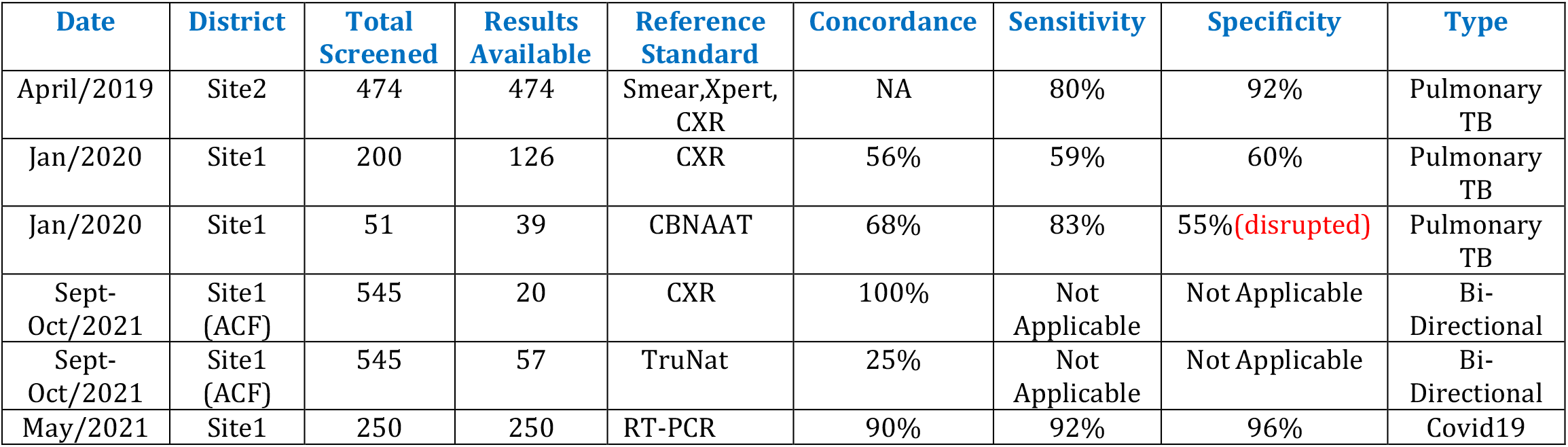

As seen from table-3, most sites reported at sensitivity between 80% - 92% when validated with CBNAAT or RT-PCR while specificity reported between 92% - 96%. While CXR is not a gold standard, the fact that there was a concordance of 56% at one site and 100% at another site makes TimBre a candidate for a screening tool at the last mile as a part of outreach programs with minimal training required by the healthcare worker.

A parallel coordinate graph plotted by Matlab Classification Learner – R2018b at screenshot-3 above depicts the solution has the ability to clearly differentiate healthy and pathological subjects while providing explainability around the Machine Learning Models for Pulmonary TB in that the pathological cases have a higher value for Spectral Centroid at frequency bands of 4000-5000 Hz. There are other variables that depict a clear demarcation such as Variance and Energy that are not depicted here. This is analogous with the Acoustic music theory of a stringed instruments under standard tuning with a low E string, when plucked at an open position that rings at a frequency of 82.3 Hz which when plucked while placing index finger on 1^st^ fret (F note) rings at 87.3 Hz at an Octave-2 for both these notes. This is analogous to accenting higher frequencies for cavitated lungs as seen from screenshot-3 above when comparing the spectral centroid for healthy (blue) and pathological (red). Another acoustic scenario mimicking the cavities would be of placing the Capodastro (Capo) at first fret and second fret respectively that now results in all strings to sound one half step and two half steps higher respectively accenting higher frequencies and different keys/scales wherein the latter case of a capo on second fret converts an Em chord into a F#m chord/key. The reason these chords are hypothesized is that a TB cough is dark & sad sounding similar to the Minor chords or vice versa. Another scenario analogous to Lung changes at high altitudes or seasonal changes could be hypothesized to a different type of tuning such as Drop-D that tunes the low E at one whole step down to D ringing at 73 Hz & the next note ringing at 78 Hz. The relative increase in frequency is identical across both the type of tunings which is validated by the standard tuning. Connecting the acoustic theory we summarize that the latent and active infection has accented higher frequencies or variance is introduced when compared to a healthy lung. Viral Pneumonia rarely causes cavities and hence the acoustic theory discussed here is focusing on TB and Lung Cancers and other conditions causing cavities.

TB infection happens in 4 stages: the initial macrophage response, the growth stage, the immune control stage, and the lung cavitation stage. These four stages happen over roughly one month (2). The first two stages can be classified as latently infected and starting third stage, Asymptomatic and Active TB can be concluded and can be mapped to the Acoustic theory described with various hypotheses. Lung cancer mimics cavity formation which however forms a single cavity that grows over a period of time & TimBre can also be used for such differential diagnosis. The formant frequencies F0, F1 & F2 for TB has no overlapping of frequencies post background noise reduction for Pulmonary TB cough sounds that is somewhat depicted by ResApp (3) in their patent at page 12 & 88, Figure 7 pertaining to overlapping frequencies but different mean, kurtosis and skewness that fits the Acoustic Guitar analogy under different conditions and the frequencies they ring under the said conditions. The overlap however is seen in healthy cough sounds that becomes a key differentiator.

## Supporting information

TB Protocol approved by IRB

CT Results for Site-2

CT Results for Site 1

## Data Availability

All baseline data corresponding to the sound files (WAV format) & feature extracted Machine Learning data is available with the founding team members of Docturnal Private Limited & is proprietary in nature. The results of the trials are summarized in this manuscript and also quoted where applicable

## Limitations

- Subjects with age <18 age are not covered and is planned as a separate trial for pediatric group. We however did not deny screening and results thereon
- Omicron related cough sounds that are predominantly URTI (upper respiratory tract infection) were not covered
- The study focused on 3^rd^ party Microphone Array - Zoom H1 & H1n models, Nokia 2.3 & Xiomi Redmi Mobile Phone Models & needs to evaluate other low cost microphones such as Bietrun

