## Supplementary material for "Acoustic Epidemiology of Pulmonary Tuberculosis (TB) & Covid19 leveraging AI/ML": TB Protocol approved by IRB

**Docturnal**

**CLINICAL STUDY PROTOCOL**

**Protocol number: TimBre-01**

**Version 1.0 dated Nov 29, 2018**

**Study title:** Multicentric study to evaluate the efficacy of TimBre software as compared to standard screening modalities in subjects clinically suspected of pulmonary tuberculosis

**Name of the modality under investigation**

TimBre Point of care device with software

**Confidential**

This document contains confidential information belonging to the sponsor. Except or otherwise written to in writing, by accepting or reviewing this document, you agree to hold this information in confidence and not copy or disclose it to others (except when required by applicable law) or use it for unauthorized purposes. In the event of actual or suspected breach of this obligation, the sponsor must be promptly notified.

| **Principal contacts** | |
| --- | --- |
| **1. Principal Investigator**  **(Site 1)** | Dr. Alben Sigamani  Group Head  Department of Clinical Research  Narayana Hrudayalaya, Bangalore |
| **2. Principal Investigator**  **(Site 2)** | Dr. AK Khan  Erragadda Chest Hospital  Hyderabad, Telengana |
| **3. Principal Investigator**  **(Site 3)** |  |
| **Sponsor details** | Mr. Rahul Pathri CEO  Docturnal Pvt Ltd, Hyderabad |

**TABLE OF CONTENTS**

1. Study synopsis	4

2. Signature page	7

3. Background and significance	8

4. Objectives	10

5. Inclusion/ exclusion criteria	11

6. Study Procedure	12

7. IRB approval and informed consent	15

8. Data collection and management	15

9. Statistical considerations	16

10. References	16

**Study synopsis**

| **Study Title:** | Multicentric study to evaluate the efficacy of TimBre software as compared to standard screening modalities in subjects clinically suspected of pulmonary tuberculosis |
| --- | --- |
| **Type of study** | Clinical evaluation of diagnostic device (with software) |
| **Study Design** | Prospective comparative multi centric non inferiority study to evaluate the effectiveness of the TimBre software when compared to the standard screening modalities in subjects presenting with symptoms suggestive of TB |
| **Objective** | **Primary Objective**  1. To assess non-inferiority of sensitivity of TimBre software to not more than 10% of the sensitivity of standard screening modalities  2. Evaluate the diagnostic performance of TimBre with standard screening modalities as measured by sensitivity, specificity, PPV and NPV  **Secondary Objective**  1. To study the influence of patient characteristics on diagnostic accuracy of TimBre software.  2. To study the efficacy of TimBre software in differentiating drug sensitive cases from drug resistant TB cases  3. To study the correlation of sputum conversion at the end of 3 months with cough characteristics  4. To study the relationship between treatment and cough characteristics using TimBre software  5. To compare the diagnostic accuracy of TimBre software between cough sample collected using Microphone array and mobile |
| **Duration of study** | 6 months (3 months for recruitment and 3 months follow up) |
| **Study population and sample size** | Study involves 2 group of participants:  **Group 1:** Participants who present with symptoms suggestive of pulmonary TB and undergoing diagnostic investigation for TB (N = 1000)  **Group 2:** Subjects non suspicious of TB (N = 500) |
| **Eligibility criteria** | **Inclusion criteria**  1. Participants of either gender above 18 years with symptoms suggestive of Pulmonary TB such as   - Cough >2 weeks, - Fever >2 weeks, - Significant weight loss (more than 4.5 kg or 5% in 6 to 12 months) - Haemoptysis, - Any abnormalities in chest radiography.   2. Who are willing to give written informed consent and comply with study related visit and procedure  **Exclusion criteria**  1. Patients with extra pulmonary TB  2. Any psychological and/or pathological condition that would interfere with study participation in the opinion of the investigator. |
| **Follow up and visits** | **No. of visits:** 2 to 3 visits (depending on sputum conversion)  Visit 1: Baseline visit.  Visit 2: Time of next sputum testing (2 months from baseline)  Visit 3: 3 months from visit 1 (only if no sputum conversion in visit 2) |
| **Study procedure** | Group 1: All eligible participants will be enrolled into the study. Details related to demographic and clinical characteristics will be collected and patient will be subjected to diagnostic test for pulmonary TB as per RNTCP guidelines. Cough sample will be obtained using the TimBre device with software. Patients who are diagnosed with TB will be started on treated as per RNTCP guidelines and followed up till the next visit for sputum testing. And cough sample will be collected again.  For group 2 participants cough sample will be collected only once along with demographic and clinical characteristics. |

**Schedule of events**

| **Study activities** | **Baseline visit (V1)** | **Follow up visit (V2)** | **Follow up visit (V3)*** |
| --- | --- | --- | --- |
| Time of visit |  | 2 months | 3 months |
| Eligibility assessment |  |  |  |
| Informed consent |  |  |  |
| Demographic data, medical history |  |  |  |
| Standard screening test |  |  |  |
| Cough sample |  |  |  |

***** only if there was no sputum conversion in visit 2

**Note:** Group 2 will have only one visit.

**Protocol signature page**

**Protocol title: Multicentric study to evaluate the efficacy of TimBre software as compared to standard screening modalities in subjects clinically suspected of pulmonary tuberculosis**

I have read this protocol and agree that it contains all necessary details for carrying out the study as described. I will conduct the study as outlined therein, including all statements regarding confidentiality. I will make a reasonable effort to complete the study within the time designated. I will provide copies of this protocol and all pertinent information to the study personnel under my supervision. I will discuss this material with them and ensure they are fully informed regarding the device and the conduct of the study.

I will conduct the study in accordance with the protocol, the Declaration of Helsinki and applicable local requirements.

Principal Investigator name and Job title: _______________________________

________________________________

Institution: ____________________________________

____________________________________

Address: ____________________________________

____________________________________

Signature: _____________________________________

Date

**1. Background**

Tuberculosis (TB) is one of the oldest known disease to affect humans caused by Mycobacterium tuberculosis usually affecting the lungs although other organs can also be involved. (1) Pulmonary tuberculosis is a global heath burden affecting predominantly low and middle income countries. TB is one of the top 10 leading causes of death and the leading cause from a single infectious agent. (2) According to WHO database 2017, 1 million children became ill with TB and another 230000 children died of TB. Death due to TB was estimated to be around 1.3 million among HIV negative individuals with an additional 3 lakhs death from HIV positive individuals (3) The disease mainly spreads through airway spread of droplet nuclei from cough.

The classical symptoms of pulmonary tuberculosis are cough with sputum (> 2 weeks), fever (> 2 weeks), malaise, weight loss, breathlessness, haemoptysis usually lasting more than 2 weeks. (4)

**Cough dynamics**

A series of experiment conducted in 1960s showed that the spread of droplet nuclei was more during cough than by talking or singing. (5) Since then there was a number of studies focusing on the cough characteristics such as cough frequency, diurnal variation etc. Proano et al (6) studied the dynamics of cough frequency in 64 adults undergoing treatment for pulmonary TB using a semi-automated ambulatory monitor Cayetano Cough Monitor. The study showed that coughs were more frequent in daytime. Two weeks of antitubercular therapy (ATT) reduced cough frequency and achieved culture conversion in one third of patients. The Cavetano Cough Monitor was validated in patients with TB and showed a sensitivity of 75.5%

Botha et al (7), studied the performance evaluation of a Tascam DR-44 WL hand held automated device in the detection of TB by analysing cough sound. The study included 396 TB positive cases and 233 healthy controls. The study showed that by adjusting the threshold for classification 95% sensitivity can be achieved with specificity of 72%

**Screening and diagnosis of TB**

Early diagnosis of pulmonary TB in an individual is crucial for TB control because it helps in early initiation of treatment as well as reduce further spread in the community. Chest X ray though useful in diagnosis has limitations due to lack of specificity and presence of atypical findings that can mimic other conditions. Definitive diagnosis depends on recovery of the organism in culture or identification using DNA or RNA amplification techniques. Traditional method of light microscopy examination for acid fast bacilli and culture remains the mainstay of diagnosis. Sputum microscopy for acid fast bacilli is rapid and cost effective but has low and variable sensitivity. Sputum culture has high sensitivity and specificity for diagnosis but takes 2 to 6 weeks for the growth of the organism. (8) Nucleic acid amplification techniques do not only help in diagnosis but also helps in identification of any resistance markers. (9)

Development of nucleic acid amplification test is considered as a significant milestone in the diagnosis and management of TB. Though it has immense benefit in diagnosing patients presenting with symptoms suggestive of TB (passive case finding), it is prohibitively expensive to be used for active case finding and in developing countries. Control of spread of TB requires a cost effective, rapid and sensitive test that can be used for active case finding.

**TimBre software application**

TimBre is a Non-Invasive & Point of Care Solution from Docturnal that screens for Pulmonary Tuberculosis Screening using a Microphone Array that is connected to a Mobile Phone Running TimBre App. The device is a XY Microphone Array that comes with a filter (pop/wind) which needs to be replaced after a positively screened result is obtained to avoid the bacilli spread to the next suspect/patient. It supports both USB & battery & an additional connector connects to Mobile Phone Jack to the Line in jack of the Microphone Array. This can also be replaced by a USB 2.0 cable. Optionally a tripod is available as needed. Attached is a document (MPA-TimBre) that provides details. (10)

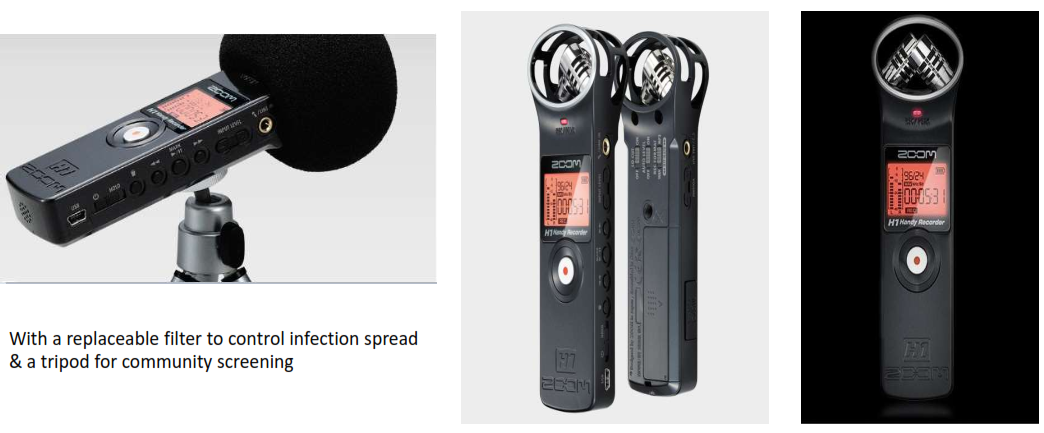

**Figure 1:** Microphone array with a replaceable filter connected.

A preliminary analytical validation study was conducted using TimBre device in 811 participants which included 300 patients with TB, 80 patients with COPD/Asthma, 100 with cough due to other reasons and healthy individuals. Results of the automatic classification (LRM) model predicted that within the data set of 811 patients, the accuracy of the model was observed to be 93% with an AUC score of 0.97, the sensitivity of 74% with the specificity of 98% (10)

**2. Objective:**

**Primary Objective**

1. To assess non-inferiority of sensitivity of TimBre software to not more than 10% of the sensitivity of standard screening modalities
2. Evaluate the diagnostic performance of TimBre with standard screening modalities as measured by sensitivity, specificity, PPV and NPV

**Secondary Objective**

1. To study the influence of patient characteristics on diagnostic accuracy of TimBre software.
2. To study the efficacy of TimBre software in differentiating drug sensitive cases from drug resistant TB cases
3. To study the correlation of sputum conversion at the end of 3 months with cough characteristics
4. To study the relationship between treatment and cough characteristics using TimBre software
5. To compare the diagnostic accuracy of TimBre software between cough sample collected using Microphone array and mobile

**3. Methodology:**

**Study design**: This is a prospective comparative multi centric non inferiority study to evaluate the effectiveness of the TimBre software when compared to the standard screening modalities in subjects presenting with symptoms suggestive of TB

**Study duration:** 6 months (3 months for recruitment and 3 months follow up)

**Study site:** The study will be conducted in 3 sites YRG hospital (Chennai), NE and NH (Bangalore)

**Population characteristics**

Study involves 2 group of participants

**Group 1:** Participants who present with symptoms suggestive of pulmonary TB and undergoing diagnostic investigation for TB (N = 1000)

**Group 2:** Subjects non suspicious of TB (N = 500)

Clinical evaluation of the software in comparison with standard screening modalities will be done using group 1 subjects.

Evaluation of false positivity of software in general population will be done in group 2 subjects.

**Clinical evaluation of the software in comparison with standard screening modalities**

**Inclusion criteria**

1. Participants of either gender above 18 years with symptoms suggestive of Pulmonary TB such as

- Cough >2 weeks,
- Fever >2 weeks,
- Significant weight loss (more than 4.5 kg or 5% in 6 to 12 months)
- Haemoptysis,
- Any abnormalities in chest radiography.

2. Who are willing to give written informed consent and comply with study related visit and procedure

**Exclusion criteria**

1. Patients with extra pulmonary TB

2. Any psychological and/or pathological condition that would interfere with study participation in the opinion of the investigator.

**Procedure**

The study will be initiated after IEC approval and registration of the protocol in the Clinical Trial registry of India.

All subjects who visit the trial site and are found to be eligible will be counselled for participation. The Principal Investigator or member of the study team will explain the trial to the subject as part of the informed consent process. Once participant has signed the consent form information such as demographic characteristics, type of presentation, comorbid illness will be collected. Participant will undergo screening test as for TB (chest X ray, 3 days sputum test for AFB or nucleic acid amplification test) as per diagnostic algorithm.

The subject will then be given instructions regarding giving cough sample. Cough sample will be collected both using Microphone array and also directly in the mobile.

**Procedure for cough sample collection**

- After connecting the device to the phone, make sure to select WAV as the format & sampling rate and bit rates are set at 44.1kHz and 16 bits respectively. Turn on low cut filter ON
- Ensure to set correct data and time on the device by simultaneously clicking the record button and the start button
- A pop filter is used prior to the coughing. (The filter will be changed for every participant)
- Start TimBre after downloading it from playstore and choose English as a language
  - Enter Hospital Name or Site Name on the first screen and ensure some mandatory fields on the next screen are entered – Mobile, Ht, Wt, Age
  - Click Next & can ignore the upload image screen till the Microphone record screen
  - At this screen, click record button and start recording the cough after clicking the red record button on the device & click it again to stop recording once the recording is completed
  - Keep the device at an arm’s length from the patients mouth while recording or while on the tripod at the time of coughing
  - Ensure the subject is wearing a surgical mask while coughing
  - <https://youtu.be/UWBfP8Gbwn0> - App video
  - Finish button ensures the transaction is completed and the data is available on the cloud for processing against the algorithms and results can be sent via an SMS or a consolidated report after each day can be sent to a hospital

**Mobile protocol**

The protocol is similar to the Microphone Array (external hardware) recording with an exception that the suspect directly onto the Mobile Phone as depicted in above screenshot. A few steps in the protocol are stated below:

1) The App is available as a private APK that is a replica of TimBre which is built using Native Android to facilitate a lossless WAV file. The patient has a window of 20 seconds to complete voluntary cough recording at their convenience after a brief introduction of their name(optional)
2) The mobile phone needs to be cleansed with a few drops of surgical spirit/cotton to avoid infection for the subsequent screenings & wait for a minimum of 3 minutes to conduct next screening to dry the device
3) User needs to wear surgical gloves and wear a n-95 mask
4) ~~Optionally,~~ The patient needs to use a Surgical Mask while coughing to avoid the spread of Bacilli

**Cough sample collection**

Cough sample will be obtained each time the patient is giving sputum sample or any other specific diagnostic test is carried out. For the confirmed PTB cases treatment will be initiated as per RNTCP guidelines. Patient will be followed up and cough sample will be collected till his treatment regimen goes from intensive phase to maintenance phase (2 to 3 months)

**No. of visits:** 2 to 3 visits (depending on sputum conversion)

Visit 1: Baseline visit.

Visit 2: Time of next sputum testing (2 months from baseline)

Visit 3: 3 months from visit 1 (only if no sputum conversion in visit 2)

**Evaluation of false positivity of software in general population**

**Group 2:** **Subjects non suspicious of TB** (N = 500)

The second group of subjects include participants presenting to hospital for other complaints. This group is included to check for false positivity of the software in the general population.

For this subjects presenting to the hospital for some other complaints and not suspicious of TB will be included. Subjects above 18 years of age of either gender willing to give written informed consent will be included for the test. The procedure will be explained to them and cough sample will be collected as mentioned earlier. Subjects who are diagnosed as positive for TB based on TimBre software will be counselled for screening test for TB as per ethical guidelines.

**Study flow**

Group-1 Subjects with symptoms suggestive of Pulmonary TB

Group 2: Subjects non suspicious of TB

V1- Eligibility assessment and Informed consent

Collection of demographic data, medical history

Standard screening test (Sputum examination, Chest X ray...)

Collection of cough sample

Follow up (**only for group 1**)

V2- At the end of 2 months

V3- At the end of 3 months (only if there was no sputum conversion in V2)

**Study activities**

| **Study activities** | **Baseline visit (V1)** | **Follow up visit (V2)** | **Follow up visit (V3)*** |
| --- | --- | --- | --- |
| Time of visit |  | 2 months | 3 months |
| Eligibility assessment |  |  |  |
| Informed consent |  |  |  |
| Demographic data, medical history |  |  |  |
| Standard screening test |  |  |  |
| Cough sample |  |  |  |

***** only if there was no sputum conversion in visit 2

**Note:** Group 2 will have only one visit.

**4. IRB approval and informed consent**

Ethics committee approval will be obtained from all the sites prior to enrolment of participants. Written informed consent will be obtained from all the study participants in the form approved by the IEC.

There is no additional risk to the subjects because of their participation in the study.

Any participant in group 2 showing positive test for TB by the software will be counselled for standard screening test.

**5. Data collection and management**

Data regarding demographic characteristics, medical history will be collected in the mobile application. An Admin portal which hosts the data from the cloud which collects the clinical, demographic and acoustic data which is processed by the proprietary algorithms and on a need basis, the results are sent to the Mobile phone used for screening as an SMS.

**6. Statistical considerations**

**A. Sample size estimation**

In a general population, the rate of detecting TB positive by either x-ray or sputum positivity among a low suspicion for TB, is 6 / 100. This can be as high as 30 – 50% in a case with clinical history.

For a non-inferiority design, keeping the proportion of positive cases being picked up by the test device and the standard test the same with a non-inferiority margin of 10%, we will need to screen 1000 suspect cases to detect nearly 60 cases with positive TB.

**B. Statistical analysis plan**

The sensitivity of TimBre software and that of standard modalities will be compared to assess non inferiority.

The primary objective of non-inferiority will be assessed by constructing a 90% confidence interval (CI) around the difference in sensitivities of TimBre and standard modalities. If The lower limit of the 90% CI is greater than -10%, then non-inferiority will be established. All the methods of validity will be computed for TimBre along with their 90% CI

10. Jha S, Pathri R, Tandon S, Reddy VG, Alfred Saxby AE. TimBre: Acoustic Based Non-Invasive Point of Care Screening of Tuberculosis. Manuscript submitted for publication.
