## Supplementary material for "Acoustic Epidemiology of Pulmonary Tuberculosis (TB) & Covid19 leveraging AI/ML": CT Results for Site-2

### STUDY STATUS REPORT

**Study title:** Multicentric study to evaluate the efficacy of TimBre software as compared to standard screening modalities in subjects clinically suspected of pulmonary tuberculosis

**Primary Objective**

1. To assess non-inferiority of sensitivity of TimBre software to not more than 10% of the sensitivity of standard screening modalities
2. Evaluate the diagnostic performance of TimBre with standard screening modalities as measured by sensitivity, specificity, PPV and NPV

**Secondary Objective**

1. To study the influence of patient characteristics on diagnostic accuracy of TimBre software.
2. To study the efficacy of TimBre software in differentiating drug sensitive cases from drug resistant TB cases
3. To study the correlation of sputum conversion at the end of 3 months with cough characteristics
4. To study the relationship between treatment and cough characteristics using TimBre software
5. To compare the diagnostic accuracy of TimBre software between cough sample collected using Microphone array and mobile

**Study group**

**Group 1:** Participants who present with symptoms suggestive of pulmonary TB and undergoing diagnostic investigation for TB (N = 1000)

**Group 2:** Subjects non suspicious of TB (N = 500) **completed**

**Group 3:** Subjects with productive cough (N= 100) - **ongoing**

**Results: Group 2 Table-1 Baseline characteristics**

| **Variables** | **Descriptive statistics (N= 430)** |
|---|---|
| Gender Male | 255 (59.3%) |
| Age | 36.0 [50.2, 28.0] |
| Height (feet) | 5.5 [5.7, 5.3] |
| Weight (kg) | 64.92 ± 12.8 |
| BMI | 24.0 [27.0, 21.0] |
| Marital Status<br>Married<br>Null<br>Single | <br>304 (70.7%)<br>13 (3.0%)<br>113 (26.3% ) |
| Smoker | 37 (8.6%) |
| Consumes Alcohol | 47 (10.9%) |
| Appetite Pattern<br>Low<br>Medium<br>High | <br>165 (38.4%)<br>255 (59.3%)<br>10 (2.3%) |

A total of 474 patients were recruited as of Feb 8, 2020. Among them 39 patients were diagnosed as positive by TimBre software. The details of their follow up are mentioned as follows:

**Table 2- Timbre results for 474 patients**

| | Number (Percentage) |
|---|---|
| Number of cases detected positive by TimBre | 39 |
| Number of cases confirmed positive | 4 (2 on treatment, one old case and one diagnosed subsequently) |
| Number of cases confirmed negative | 10 |
| Number of cases lost to follow up | 13 |
| Number of participants who were symptom free on telephonic follow up | 12 |

**Number of cases missed diagnosis of TB by TimBre 1**

For a total of 474 participants screened 39 were detected as positive by TimBre software. Among the 39 participants 4 were positive. Two were on ATT at the time of testing for TimBre software. One was an old case with no active bacilli in sputum. One patient was later found to be positive on further evaluation.

Ten participants had clinical evaluation and was confirmed as negative for TB by either clinical examination or sputum testing. Thirteen patients could not be reached further for telephonic follow up or hospital visit. Remaining 12 patients were followed telephonically for symptoms suggestive of TB like chronic cough, fever and weight loss. None of these 12 patients were symptomatically positive for tuberculosis.

One patient who was positive by GenXpert was missed by the software.

**Table 3 Sensitivity, specificity of TimBre**

| | **Clinical diagnosis positive** | **Clinical diagnosis negative** | **Total** |
|---|---|---|---|
| **TimBre diagnosis positive** | **4** | **35** | **39** |
| **TimBre diagnosis negative** | **1** | **434** | **435** |

| **Statistic** | **Value** | **95% CI** |
|---|---|---|
| Sensitivity | 80.00% | 28.36% to 99.49% |
| Specificity | 92.54% | 89.77% to 94.75% |
| Positive Likelihood Ratio | 10.72 | 6.24 to 18.43 |
| Negative Likelihood Ratio | 0.22 | 0.04 to 1.25 |
| Disease prevalence (*) | 1.05% | 0.34% to 2.44% |
| Positive Predictive Value (*) | 10.26% | 6.23% to 16.42% |
| Negative Predictive Value (*) | 99.77% | 98.69% to 99.96% |
