## Supplementary material for "Acoustic Epidemiology of Pulmonary Tuberculosis (TB) & Covid19 leveraging AI/ML": CT Results for Site 1

### Interim report- Telangana state

**Study title:** Multicentric study to evaluate the efficacy of TimBre software as compared to standard screening modalities in subjects clinically suspected of pulmonary tuberculosis

**Sponsor details:**

**Name**

**Address**

**Site details:**

**Site 1**

**Principal Investigator:**

**Address:**

**Site 2**

**Principal Investigator:**

**Address:**

**Primary Objective**

1. To assess non-inferiority of sensitivity of TimBre software to not more than 10% of the sensitivity of standard screening modalities
2. Evaluate the diagnostic performance of TimBre with standard screening modalities as measured by sensitivity, specificity, PPV and NPV

**Secondary Objective**

1. To study the influence of patient characteristics on diagnostic accuracy of TimBre software.
2. To study the efficacy of TimBre software in differentiating drug sensitive cases from drug resistant TB cases
3. To study the correlation of sputum conversion at the end of 3 months with cough characteristics
4. To study the relationship between treatment and cough characteristics using TimBre software
5. To compare the diagnostic accuracy of TimBre software between cough sample collected using Microphone array and mobile

A multi-centre software evaluation study was proposed with the above-mentioned objectives. Patients presenting with symptoms suspicious of TB were enrolled from 2 sites (Warangal and Gadwal) Patients underwent standard diagnostic testing for tuberculosis followed by testing using TimBre device.

A total of 1000 patients are planned to be enrolled with 28 patients in Warangal site and 132 patients in Gadwal district enrolled so far. Recruitment was on hold in Warangal on account of the pandemic. Interim analysis of data from Gadwal site is presented below. Mean age of the study population was 51.41 ± 15.64 with 67% male. Prevalence of TB in the study population was 58.4%. Preliminary analysis shows a sensitivity of 58.90% (46.77% to 70.29%) and specificity 59.62% (45.10% to 72.99%) for the TimBre software. Detailed analysis regarding the accuracy of TimBre software in the screening of Tuberculosis will be presented on completion of data collection.

#### List of tables

**Site-1 Warangal district**

**Table-1 Baseline demographic characteristics**

| **Variable name** | **Value (N= 25)** |
|---|---|
| Age | 51.12 ± 15.38 |
| Male | 17 (65.4%) |
| Height | 5.54 ± 0.39 |
| Weight | 47.06 ± 11.22 |
| BMI | 16.83 ± 4.21 |

**Table-2 Comparison of Timbre results with CBNAAT**

| | Disease | No disease |
|---|---|---|
| Timbre positive | 5 | 10 |
| Timbre negative | 1 | 10 |

**Table-3 Diagnostic accuracy of TimBre**

| | |
|---|---|
| Total number | 26 |
| Sensitivity | 0.83 (0.43 to 0.97) |
| Specificity | 0.50 (0.29 to 0.70) |
| PPV | 0.33 (0.15 to 0.58) |
| NPV | 0.90 (0.62 to 0.98) |
| Prevalence | 0.23 |

#### Site-2 Gadwal district

**Table-4 Baseline demographic characteristics**

| **Variable name** | **Value (N= 125)** |
|---|---|
| Age | 51.41 ± 15.64 |
| Male | 83 (66.93%) |
| Height | 5.20 ± 0.34 |
| Weight | 45.78 ± 10.81 |
| BMI | 18.70 ± 4.15 |
| Smoking | 21 (16.93%) |
| Appetite<br>Low<br>Medium | <br>89 (71.77%)<br>35 (28.22%) |
| Alcohol | 23 (18.54%) |

**Table-5 Comparison of TimBre results with chest Xray**

| | Disease | No disease | |
|---|---|---|---|
| Timbre positive | 43 | 21 | 64 |
| Timbre negative | 30 | 31 | 61 |
| | 73 | 52 | |

**Table-6 Diagnostic accuracy of TimBre**

| | | |
|---|---|---|
| Total number | 125 | |
| Sensitivity | 58.90% | (46.77% to 70.29%) |
| Specificity | 59.62% | (45.10% to 72.99%) |
| PPV | 67.19% | (58.29% to 75.00%) |
| NPV | 50.82% | (42.03% to 59.56%) |
| Prevalence | 58.40% | (49.25% to 67.15%) |
